# High viral loads: what drives fatal cases of COVID-19 in vaccinees? – an autopsy study

**DOI:** 10.1101/2021.12.03.21267155

**Authors:** Klaus Hirschbühl, Tina Schaller, Bruno Märkl, Rainer Claus, Eva Sipos, Lukas Rentschler, Andrea Maccagno, Bianca Grosser, Elisabeth Kling, Michael Neidig, Thomas Kröncke, Oliver Spring, Georg Braun, Hans Bösmüller, Maximilian Seidl, Irene Esposito, Jessica Pablik, Julia Hilsenbeck, Peter Boor, Martin Beer, Sebastian Dintner, Claudia Wylezich

## Abstract

**Background:** The rate of SARS-CoV-2 breakthrough infections in vaccinees is becoming an increasingly serious issue.

**Objective:** To determine the causes of death, histological organ alteration, and viral spread in relation to demographic, clinical-pathological, viral variants, and vaccine types.

**Design:** Comprehensive retrospective observational cohort study. Setting: Consecutive cases from four German academic medical centers.

**Patients:** Deceased with proven SARS-CoV-2 infection after vaccination who died between January and November 2021. Collections of 29 vaccinees which were analyzed and compared to 141 nonvaccinated control cases.

**Results:** Autopsies were performed on 16 partially and 13 fully vaccinated individuals. Most patients were elderly and suffered from several relevant comorbidities. Real-time RT-PCR (RT-qPCR) identified a significantly increased rate of generalized viral dissemination within the organism in vaccinated cases versus nonvaccinated cases (45% vs. 16%, respectively; P = 0.008). Vaccinated cases also showed high viral loads, reaching Ct values below 10, especially in the upper airways and lungs. This was accompanied by high rates of pulmonal bacterial or mycotic superinfections and the occurrence of immunocompromising factors such as malignancies, immunosuppressive drug intake, or decreased immunoglobulin levels. All these findings were particularly accentuated in partially vaccinated patients compared to fully vaccinated individuals. A fatal course after vaccination occurred in only 14% of all COVID-19 deceased in Augsburg.

**Limitations:** Restricted number of cases

**Conclusions:** Fatal cases of COVID-19 in vaccinees were rare and often associated with severe comorbidities or other immunosuppressive conditions. Interestingly, we observed striking virus dissemination in our case study, which may indicate a decreased ability to eliminate the virus in patients with an impaired immune system. However, the potential role of antibody-dependent enhancement must also be ruled out in future studies.

**Funding source:** This work was supported by the German Registry of COVID-19 Autopsies (www.DeRegCOVID.ukaachen.de) and funded by the Federal Ministry of Health (ZMVI1-2520COR201), the Federal Ministry of Education and Research within the framework of the network of university medicine (DEFEAT PANDEMICs, 01KX2021), and the German Federal Ministry of Food and Agriculture through the Federal Office for Agriculture and Food (project ZooSeq, grant number 2819114019).

---

Reaching herd immunity against SARS-Coronavirus-2 (SARS-CoV-2) by recovering from an infection is not reasonable given the limitations of health systems around the world. Therefore, vaccination remains, at the moment, the only option for coping with the pandemic (1). Vaccines against SARS-CoV-2 targeting the viral spike protein have been available since the end of 2020. In Europe, four vaccines (BNT162b2, mRNA-1273, AZD1222, and Ad26.COV2.S), which have demonstrated efficacy of up to 95% against COVID-19, have been approved by the European Medicines Agency (EMA) and put into use (2-5). Next to protection from infections, avoiding severe clinical courses is the main goal of vaccination against SARS-CoV-2. It is anticipated that infection and disease due to SARS-CoV-2 infection may occur despite vaccination, even after the vaccination scheme is completed (6). With regard to the reason for those infections despite completed vaccination, a distinction must be drawn between “vaccination failure” and “breakthrough infections”. Vaccination failure is usually defined as the failure of the immune system to build effective protection by antibody- and T-cell-based responses against a virus. In contrast, breakthrough infections occur, although the antibody titers achieve sufficient values (7, 8). On 28 October 2021, 1078 such infections with fatal outcomes were recorded in Germany (9). Another aspect is the protective potential of a partial vaccination after the application of the first dose and the role of different variants of SARS-CoV-2. In a large trial, the efficacy of a single dose of Ad26.COV2.S against moderate and severe COVID-19 was 52% and 64%, respectively, indicating fast immunization in a broad part of the population (4). Other studies have shown comparable results (10-12). A certain degree of immune escape of so-called variants of concern (VOC), e.g. for the beta variant, has been described by several authors for different vaccines (13-16).

During the COVID-19 pandemic, autopsies gained remarkable importance for understanding the pathophysiology of the new disease. In particular, the viral effects on different organs in severe and lethal cases can, in most cases, be investigated only by thoroughly performed autopsies in concert with sophisticated, state-of-the-art diagnostic methods (17, 18). Most relevant COVID-19-associated organ alterations, such as diffuse alveolar damage, endotheliitis, and the role of thromboembolic events, have been described based on autoptic results (19-30). However, despite this high autoptic activity, especially in Europe and the U.S., reports from autopsies of SARS-CoV-2 breakthrough infections are widely lacking. Currently, only a single case report from Germany of a partially vaccinated case is available (31).

This multicenter retrospective study aimed to provide data from a series of fatal cases of COVID-19 after partial and full vaccination. Special attention was paid to the identification of risk factors, the direct causes of death, and viral dissemination.

## METHODS

### Case Collections

The cases of this study group were collected between the end of January and October 2021. Twenty-three of the 29 autopsies were carried out at the University Medical Center of Augsburg. The six remaining cases were included from the University Medical Centers of Düsseldorf (3), Dresden (2), and Tübingen (1). Importantly, “full vaccination” was defined as receiving two doses of the vaccine, with the second dose at least 14 days before the onset of symptoms. All autopsies of these cases were performed at the University Hospital Augsburg. All cases that did not fulfill these criteria were classified as “partially vaccinated.” All cases outside Augsburg belong to this group. According to known risk factors for a severe course of COVID-19 (e.g., cardiovascular diseases, diabetes, lung diseases, obesity, cancer, older age, immunosuppression), all but one of 16 partially vaccinated and all 13 fully vaccinated cases had at least one of the relevant comorbidities. The demographic data, together with clinical-pathological data, are provided in Figure 1 and Tables 1 and 2.

**Table 1.**
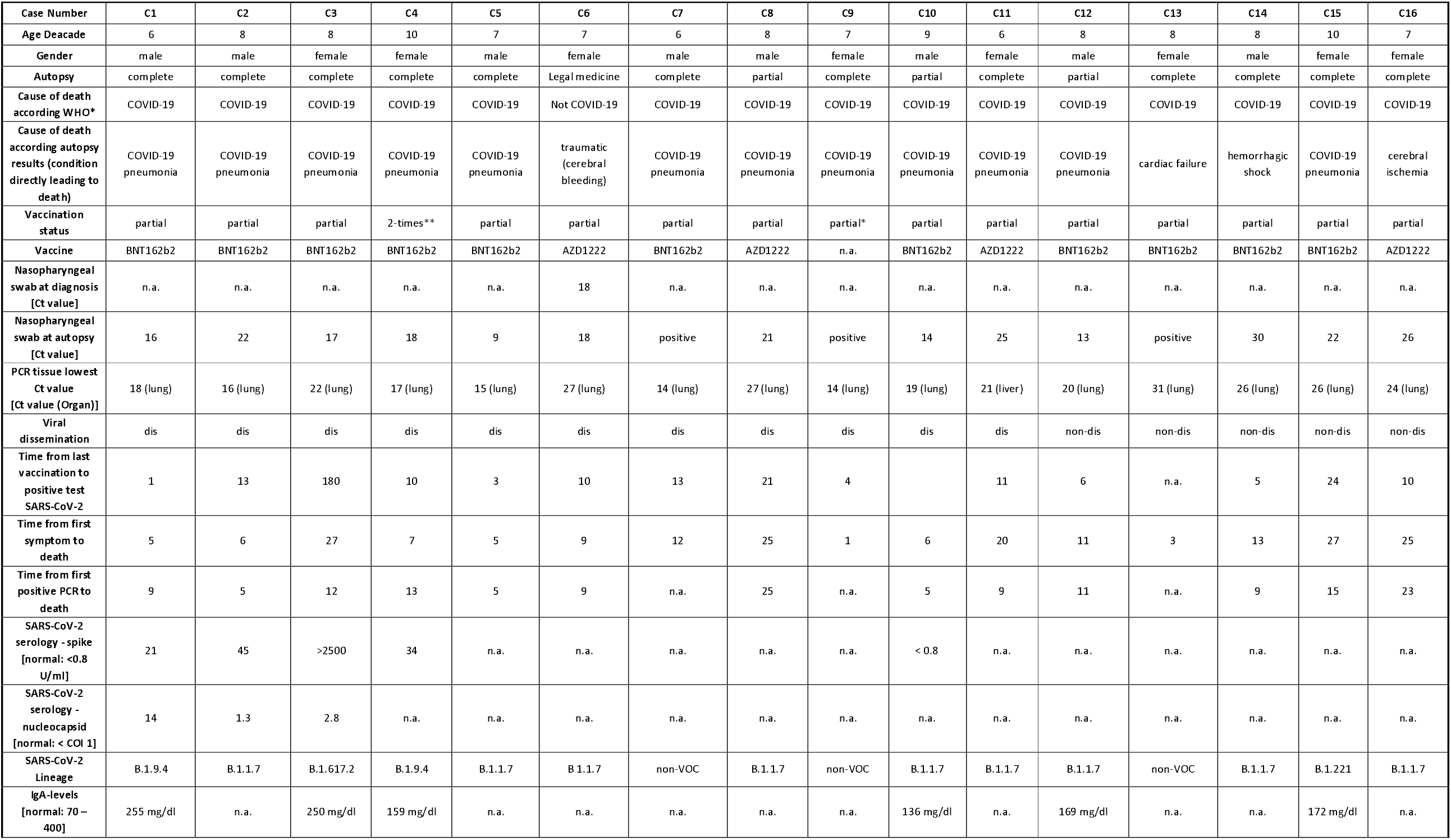
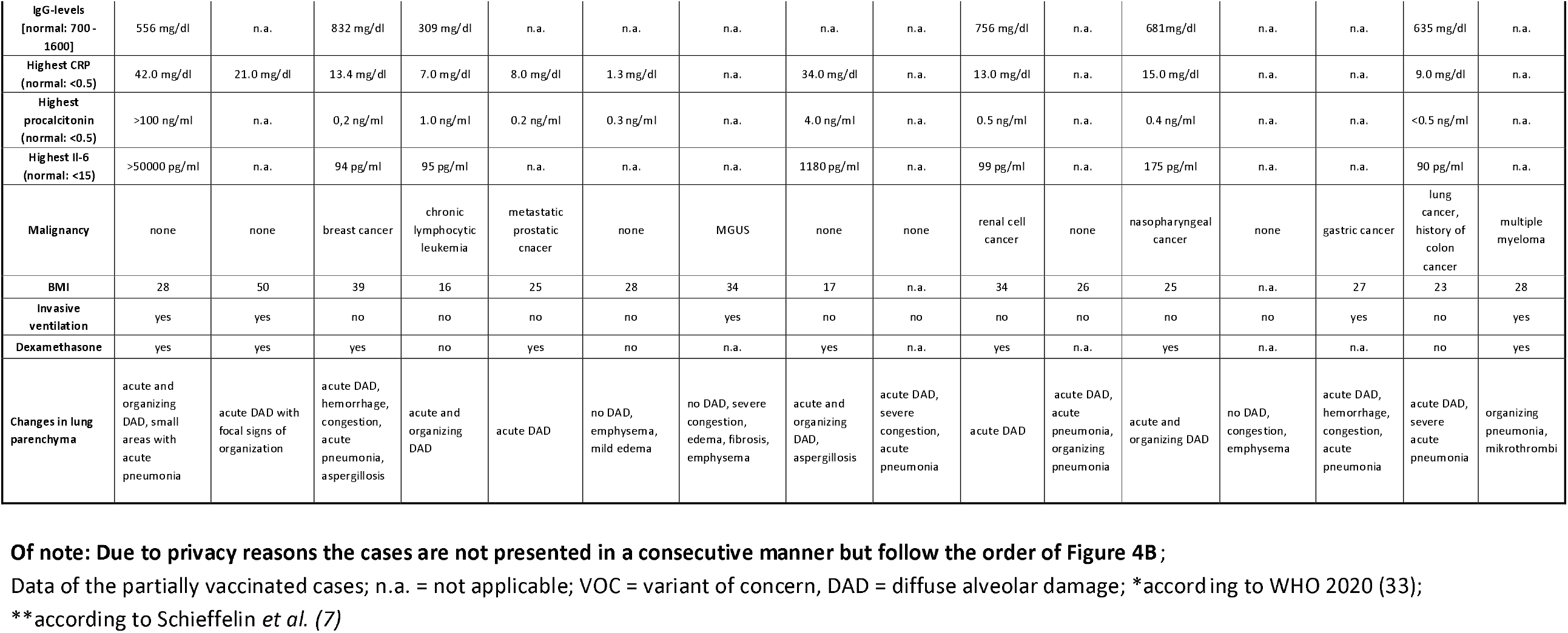

**Table 2.**

| Case Number | C17 | C18 | C19 | C20 | C21 | C22 | C23 | C24 | C25 | C26 | C27 | C28 | C29 |
| --- | --- | --- | --- | --- | --- | --- | --- | --- | --- | --- | --- | --- | --- |
| Age Decade | 8 | 9 | 9 | 9 | 8 | 8 | 8 | 7 | 7 | 8 | 8 | 6 | 10 |
| Gender | male | male | female | female | female | male | male | female | male | male | male | male | female |
| Autopsy | complete | complete | partial | partial | partial | complete | partial | complete | complete | partial | complete | complete | complete |
| Cause of death according WHO* | COVID-19 | COVID-19 | COVID-19 | COVID-19 | COVID-19 | COVID-19 | COVID-19 | COVID-19 | COVID-19 | COVID-19 | COVID-19 | COVID-19 | not COVID-19 |
| Cause of death according autopsy results (condition directly leading to death) | COVID-19 pneumonia and myocardial infarction | COVID-19 pneumonia and cardiac failure | respiratory failure** | COVID-19 pneumonia | Myocardial infarction or pulmonary embolism C19 associated | sepsis | COVID-19 pneumonia | aspiration pneumonia | COVID-19 pneumonia | COVID-19 pneumonia | COVID-19 pneumonia | COVID-19 pneumonia | myocardial infarction and nephric abscess |
| Type of infection*** | breakthrough | vaccination failure | n. d. | breakthrough | breakthrough | breakthrough | vaccination failure | n. d. | breakthrough | breakthrough | breakthrough | breakthrough | asymptomatic infection |
| Vaccination status | complete | complete | complete | complete | complete | complete | complete | complete | complete | complete | complete | complete | complete |
| Vaccine | BNT162b2 | BNT162b2 | AZD1222 | BNT162b2 | BNT162b3 | BNT162b2 | Sinovac | BNT162b2 | BNT162b2 | BNT162b2 | BNT162b2 | BNT162b2 | BNT162b2 |
| PCR nasopharyngeal swab at diagnosis [Ct value] | n.a. | 30.3 | n.a. | 34.5 | 13 | n.a. | n.a. | n.a. | n.a. | n.a. | n.a. | 22 | 41 |
| PCR nasopharyngeal swab at autopsy [Ct value] | 14 | 13 | 14 | 18 | 10 | n.a.**** | n.a. | 11 | neg***** | 34 | 20 | 18 | 40 |
| PCR tissue lowest Ct value [Ct value (organ)] | 18 (lung) | 17 (lung) | 20 (lung) | 21 (lung) | 25 (lung) | 27 (lung) | 27 (lung) | 23 (lung) | 17 (lung) | neg | 20 (lung) | 27 (lung) | neg. |
| Viral dissemination | dis | dis | dis | dis | dis # | non-dis | non-dis | non-dis | non-dis ## | non-dis | non-dis | non-dis | non-dis ### |
| Time from last vaccination to positive test SARS-CoV-2 | n.a. | 249 | 58 | 283 | 150 | 225 | 28 | 105 | 100 | 150 | 140 | 140 | 120 |
| Time from first symptom to death | 12 | 5 | 11 | 8 | 2 | 10 | 16 | 5 | 16 | 24 | 18 | 21 | 4 |
| Time from first positive PCR to death | 4 | 5 | 10 | 9 | 2 | 9 | 13 | 2 | 6 | 20 | 18 | 16 | 1 |
| SARS-CoV-2 serology - spike [normal: <0.8 U/ml] | 407 | <0.8 | n.a. | >2500 | 278 | >2500 | <0.8 | n.a. | >2500 | 223 | >2500 | 154 | >2500 |
| SARS-CoV-2 serology - nucleocapsid [normal: < COI 1] | neg. | neg. | n.a. | neg. | neg. | neg. | 2.89 | n.a. | neg. | 33 | 21.6 | 11.1 | 120 |
| SARS-CoV-2 lineage | B.1.617.2 | B.1.617.2 | B.1.1.7 | B.1.617.2 | B.1.617.2 | B.1.617.2 | B.1.617.2 | B.1.1.7 | B.1.617.2 | B.1.617.2 | B.1.617.2 | B.1.617.2 | B.1.1.7 |
| IgA-levels [normal: 70 – 400] | n.a. | n.a. | 98 mg/dl | n.a. | n.a. | 58 mg/dl | n.a. | n.a. | n.a. | 171 mg/dl | n.a. | n.a. | n.a. |
| IgG-levels [normal: 700 - 1600] | n.a. | n.a. | 511 mg/dl | n.a. | n.a. | 364 mg/dl | n.a. | n.a. | n.a. | 820 mg/dl | n.a. | n.a. | n.a. |
| Highest CRP (normal: <0.5) | 35.0 mg/dl | 17.7 mg/dl | 12.0 mg/dl | 7.4 mg/dl | 1.9 mg/dl | 19.9 mg/dl | 26.5 mg/dl | n.a. | 34.4 mg/dl | 34.2 mg/dl | 26.0 mg/dl | 23.8 mg/dl | 20.6 mg/dl |
| Highest procalcitonin (normal: <0.5) | 50.0 ng/ml | n.a. | 0.16 ng/ml | 0.1 ng/ml | n.a. | 0.9 ng/ml | 2.1 ng/ml | n.a. | 2.2 ng/ml | 2.2 ng/ml | 21.0 ng/ml | 0.7 ng/ml | >100 ng/ml |
| Highest IL-6 (normal: <15) | 914 pg/ml | n.a. | 16 pg/ml | n.a. | n.a. | 196 pg/ml | >50.000 pg/ml | n.a. | 1720 pg/ml | 534 pg/dl | 477 pg/dl | 283 pg/ml | n.a. |
| Malignancy | none | none | MPN | none | none | none | none | none | none | none | prostate cancer | none | breast cancer |

| BMI | 37 | 25 | 24 | 35 | 25 | 16 | 35 | n.a. | 16 | 31 | 28 | 27 | 25 |
| --- | --- | --- | --- | --- | --- | --- | --- | --- | --- | --- | --- | --- | --- |
| Invasive ventilation | no | no | no | no | no | no | yes | no | yes | yes | yes | yes | no |
| Dexamethasone | yes | no | no | yes | no | no | yes | no | yes | yes | yes | yes | no |
| Changes in lung parenchyma | acute DAD | mild acute DAD, acute pneumonia, aspergillosis, severe emphysema, severe congestion | no DAD, congestion of blood vessels | mild acute DAD and acute pneumonia | mild unspecific alterations, no DAD | acute pneumonia, very mild acute DAD, marked mixed pneumoconiosis | mild acute DAD, congestion | emphysema, acute pneumonia | acute/organizing DAD severe emphysema, acute pneumonia | moderate acute DAD | acute/organizing DAD | organizing DAD with residual acute DAD, aspergillosis | UIP, no DAD |

Data of the fully vaccinated cases.
**Of note: Due to privacy reasons the cases are not presented in a consecutive manner but follow the order of Figure 4B;**
n.d. = not done; n.a. = not applicable; dis = dissemination; DAD = diffuse alveolar damage; UIP = usual interstitial pneumonia; VOC = variant of concern. \*
according to WHO 2020 (33); \*\* only mild changes in lungs, no proven other cause of death, but only partial autopsy; \*\*\* according to Schieffelin *et al.* (7);
\*\*\*\* tracheal: 17; \*\*\*\*\* tracheal: 14; # only three values, but soft tissue positive; ## but cerebrospinal fluid positive; ### only nasopharyngeal swab positive

**Figure 1:**
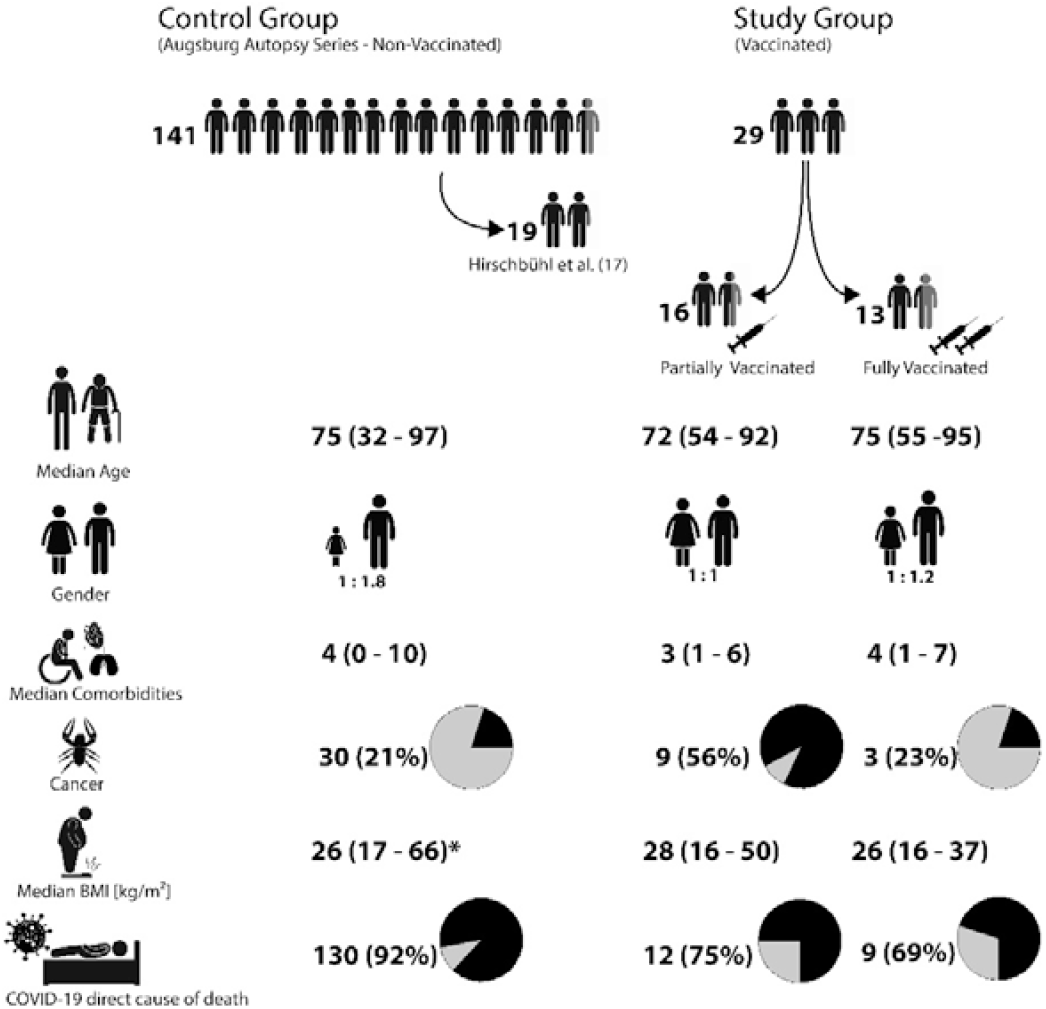
The different study and control groups of the study and basic clinical data. BMI = body mass index; * n =102

The type of infection, breakthrough versus vaccination failure, was set according to the definition described by Schieffelin et al. (7). Breakthrough infection was defined as a symptomatic lower respiratory tract infection in a case with at least a low response to full vaccination. Nonvaccinated cases of the Augsburg autopsy series served as controls (n = 141). Nineteen of these cases were published previously, providing data regarding the individual viral spread in fatal cases of COVID-19 (17). The time course of patient numbers and autopsies is given in Figure 2. Written consent was obtained from the next of kin. This study was approved by the internal review board of the medical center-Augsburg (BKF No. 2020–18) and the ethics committee of the University of Munich (Project number 20–426, COVID-19 registry of the University hospital Augsburg, the ethics committee of University Dresden (BO-EK-175052020), the ethics committee of University Düsseldorf (2020-971), and the ethics committee of University Tübingen (236/2021BO2).

**Figure 2.**
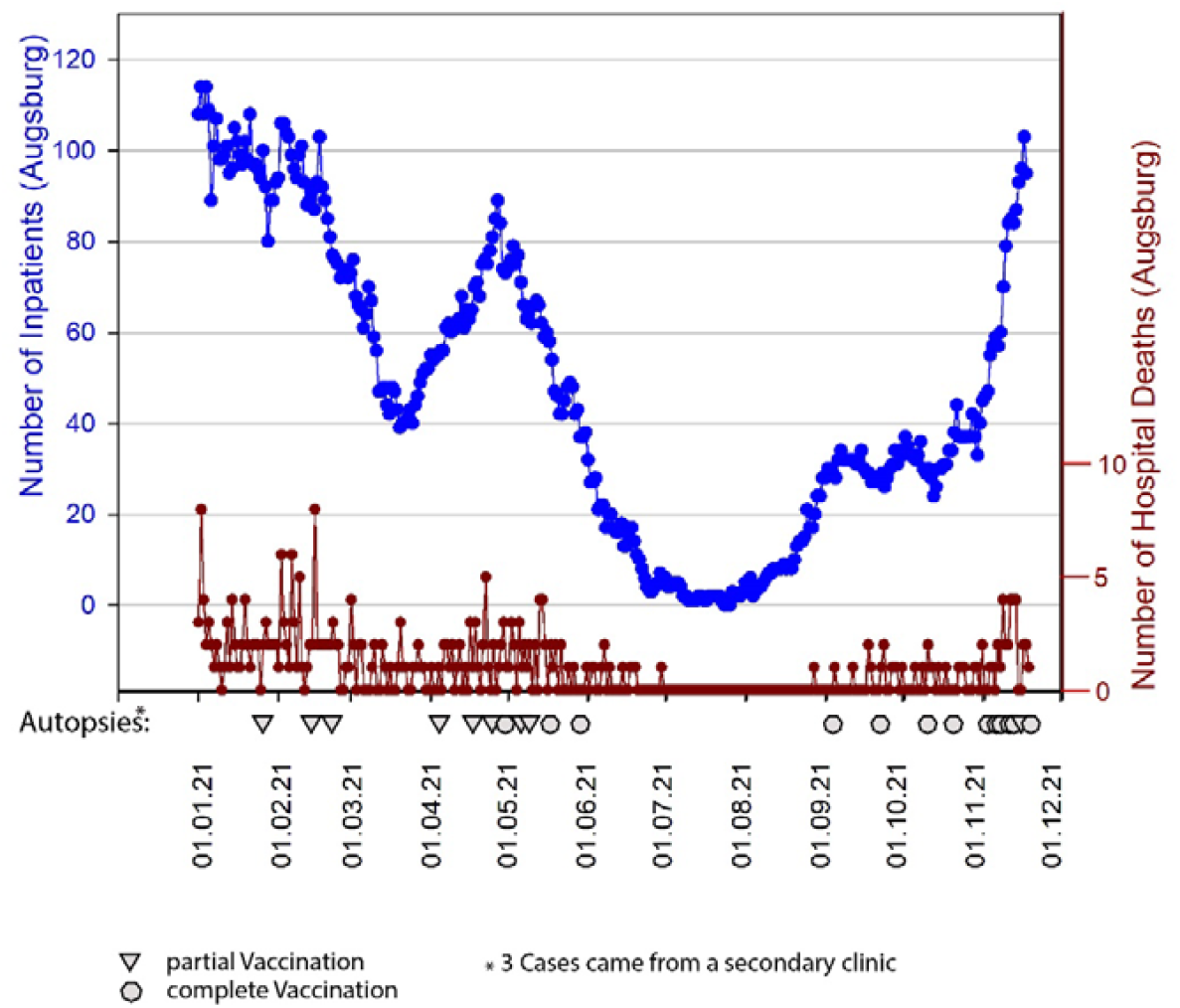
COVID-19 inpatients from 01-2021to 11-2021(blue line) and number of COVID-19 deceased during this period (brown line). Triangles and circles indicate autopsies of vaccinated deceased.

### Autopsy, Sample Collection, and Histology

The techniques of autopsy and histology workup have been described previously (17). Depending on the consent of the relatives, complete autopsies with the opening of all body cavities or partial autopsies of differing extent were performed. In case of partial autopsy, tissue samples of the thoracic and abdominal organs were obtained from epigastric access or using a recently established scopic technique (32). Regardless of the autopsy technique, tissue samples from organs and soft parts were collected and fixed in 10% buffered formalin for at least 24 hours, and then embedded in paraffin. Liquid samples, if available, were collected from cerebrospinal fluids and effusions. In cases at the Augsburg Center, nasopharyngeal swabs were also performed. This was done immediately before performing the autopsy.

The causes of death were determined according to the official definition of the WHO (33), with an indication of the disease underlying the death. Additionally, the immediate cause of death was determined.

### Real-time RT-PCR (RT-qPCR)

The RT-qPCR method used has also recently been described (17). RNA was extracted from FFPE sections and swabs using the Promega Maxwell automatic purification system (Promega Corporation, Madison, WI, USA). RT-qPCR was performed on a QuantStudio 5 Dx real-time PCR Instrument (Thermo Fisher, Carlsbad, CA, USA) using the Taq-Path COVID-19 CE-IVD RT-PCR Kit (Thermo Fisher, Pleasanton, TX, USA). The cycle threshold (Ct) values were classified in six categories (<10; 11–17; 18–24; 25–29; 30–40; negative). In cases where whole genome sequencing of the virus was not available, the variants were determined by PCR-based mutation analysis. Importantly, because of its essential role in this study, viral dissemination was defined in complete autopsies as RT-qPCR viral RNA detection in at least six of seven locations (lung, heart, central vessel, kidney, liver, spleen, and mediastinal fat). Due to the limited availability of samples from different organs, the definition had to be adapted in partial autopsies. In these situations, the PCR positivity of all samples was assumed to be dissemination.

### RNA In Situ Hybridization

RNAscope in situ hybridization (ISH) assays were conducted at the Department of Pathology of the University Hospital of Augsburg to detect SARS-CoV-2 genomic RNA in FFPE tissues. The analysis was restricted to lung samples with RT-qPCR Ct-values of < 25. ISH was performed using SARS-CoV-2 RNA-specific antisense probes designed and synthesized by Advanced Cell Diagnostics (ACD, Palo Alto, CA, USA; Cat. No: 848568). Probes specific to the dihydrodipicolinate reductase B mRNA of *Bacillus subtilis* (DapB) and peptidylprolyl isomerase B (Hs-PPIB) (ACD, Cat. No: 313908) or ubiquitin C (Hs-UBC) (ACD, Cat. No: 312028) were used as negative and positive controls, respectively, to assess assay specificity and RNA integrity. The RNAscope ISH assays were conducted using the RNAscope 2.5 LS Reagent kit-BROWN (ACD, Cat. No: 322100) on the Leica BOND-RX System (Leica, Germany), according to the automated RNAscope protocol optimized for use on this platform. FFPE sections were baked and deparaffinized in the instrument, followed by target retrieval for 25 min at 95 °C in 1X target retrieval solution and protease treatment for 35 min at 40 °C. Subsequently, slides were incubated with the ready-to-use (RTU) target probe mixtures for 2 h at 42 °C, followed by signal amplification with a set of specific amplifiers (AMP1-6). Chromogen detection and hematoxylin counterstaining were performed using a bond polymer refine detection kit (Leica, Cat. No.: DS9800) on the Leica BOND (Leica, Wetzlar, Germany).

### SARS-CoV-2 Sequencing and Sequence Analysis

The SARS-CoV-2 viral target genome amplicon libraries were constructed using the QIAseq SARS-CoV-2 Primer Panel V1 (Qiagen, Germany), coupled with the QIAseq FX DNA library kit (Qiagen, Germany), following the manufacturer’s protocols. Briefly, 5 μl of total RNA of swab samples of different viral inputs (Ct value between 18 and 28) was reverse transcribed to synthesize cDNA using random hexamers. Then, 5 μl of cDNA was evenly split into two PCR pools (2.5 μl for each pool) and amplified into 400 bp amplicons using two sets of primers that cover 99% of the entire SARS-CoV-2 genome. The primer panel was designed based on ARTIC V3 primers. PCR was performed according to the manufacturer’s instructions with 35-cycle amplification. After amplification, the contents of the two PCR pools were combined into one single tube for each sample, followed by an AMPure bead cleanup, following the manufacturer’s instructions. The purified amplicons were quantified using the Quantus System (Promega) and normalized for DNA library construction. Enzymatic fragmentation and end repair were performed to generate 250 bp DNA fragments. The fragmentation time was set to 20 min. The AMPure bead cleaned-up DNA libraries were further amplified, i.e., 8 cycles for the 40 ng input of amplicons or 20 cycles for the 1.8 ng input of amplicons. The final libraries were quantified by Quantus (Promega) prior to sequencing. Next, the libraries were multiplexed with different barcodes and pooled at 2 nM in equimolar amounts. The pooled libraries were clustered and sequenced on an Illumina MiSeq V2 flow cell at a final concentration of 9 pM (Illumina, Inc., San Diego, CA, USA).

The SARS-CoV-2 whole genome sequence of some cases was generated using the application of a generic metagenomics workflow (34) in combination with a capture enrichment procedure using myBaits (35) or the Ion AmpliSeq SARS⍰CoV⍰2 Research Panel (ThermoFisher) with 10 μg RNA as input. For the latter application, an Ion Chef instrument was used. After a quality check and quantification (33), the libraries were pooled and sequenced on an Ion Torrent S5XL instrument (ThermoFisher).

For analysis after sequencing of each library, FASTQ files were imported into CLC Genomics Workbench version 21.0.1 (Qiagen A/S, Vedbæk, Denmark) with the CLC SARS-CoV-2 workflow. Briefly, reads were imported, trimmed, and mapped to the SARS-CoV-2 reference sequence Wuhan-Hu-1 (MN908947.3). Alternatively, raw data sets were analyzed using the Genome Sequencer Software Suite (version 2.6; Roche, Mannheim, Germany https://roche.com), with default software settings for quality filtering and mapping, and using the reference mentioned above. The SARS-CoV-2 genome sequences generated in this study are available under ENA study accession number PRJEB49094.

### Phylogenetic Analysis of SARS-CoV-2 Sequences

Sequences were attributed to SARS-CoV-2 lineages using pangolin (https://pangolin.cog-uk.io/, (36)). In addition, the obtained SARS-CoV-2 genome sequences were aligned together and with sequences retrieved from GenBank using MAFFT version 7.388 (37) as implemented in Geneious version 10.2.3 (Biomatters, Auckland, New Zealand). Phylogenetic trees were constructed using PhyML version 3.0 (38), using the GTR + GAMMA + I model with 100 bootstrap replications, and MrBayes version 3.2.6 (39), using the GTR model with eight rate categories and a proportion of invariable sites in the Geneious software package. The Bayesian analysis was performed for 1,000,000 generations and sampled every 1000 generations for four simultaneous chains.

### Statistics

Depending on group size, categoric data were compared either with Chi-Square or the Fisher’s Exact test. For the comparison of continuous data, the Student’s t-test was used for normally distributed data. Ranked data were compared using the Mann-Whitney Rank Sum test or analysis of variance (ANOVA) on Ranks test. A P-value of less than 0.05 was considered significant. All tests were performed using the Sigma Plot software package 13.0 (Systat, San Jose, CA, USA).

## RESULTS

### Partially Vaccinated Cases

Partial vaccination was present in 16 cases. The median time between vaccination and the first positive PCR test was 10 days (range: 1–24). Eleven patients received the BNT162b2 vaccine (BioNTech), and four were vaccinated with AZD1222 (AstraZeneca). In one case, information regarding the vaccine was not available. There was a nonsignificant trend toward the predominance of females among AZD1222 vaccinated patients (P = 0.282). Otherwise, the female and male genders were balanced in this group. No correlation was found between the vaccine and other clinical-pathological data. The viral variants included six cases of non-VOCs (B.1.221, B.1.9.4) and ten VOCs (nine alpha, one delta; see Table 1 and for lineage assignment). This finding reflected the prevalent variants at the respective times of the pandemic (Figures 2 and 4).

In one case (C6), the SARS-CoV-2 infection was most likely not the cause of death, according to the definition of the WHO. This patient died due to traumatic cerebral bleeding. In the remaining 15 cases, the underlying cause of death in terms of the WHO definition (33) was COVID-19. The direct cause of death in three of the 15 cases was cerebral ischemia, cardiac failure, and bleeding, while 12 patients died directly due to severe COVID-19 pneumonia with diffuse alveolar damage (DAD) (Table 1). The histological presentation was similar to that of nonvaccinated cases (Figure 3). RT-qPCR-based detection of SARS-CoV-2 RNA from upper airway swabs revealed low Ct values (median: 18; range: 9– 30), indicating high viral loads. Moreover, remarkable intraindividual viral dissemination was identified in 11 out of 16 cases (Figure 4). This rate was higher than in fully vaccinated cases, with a rate of 38% (P = 0.144), but failed significance. However, it was significantly higher compared to the 19 previously published nonvaccinated cases of the first wave (17), with 16% (P = 0.002) showing such a dissemination pattern.

**Figure 3.**
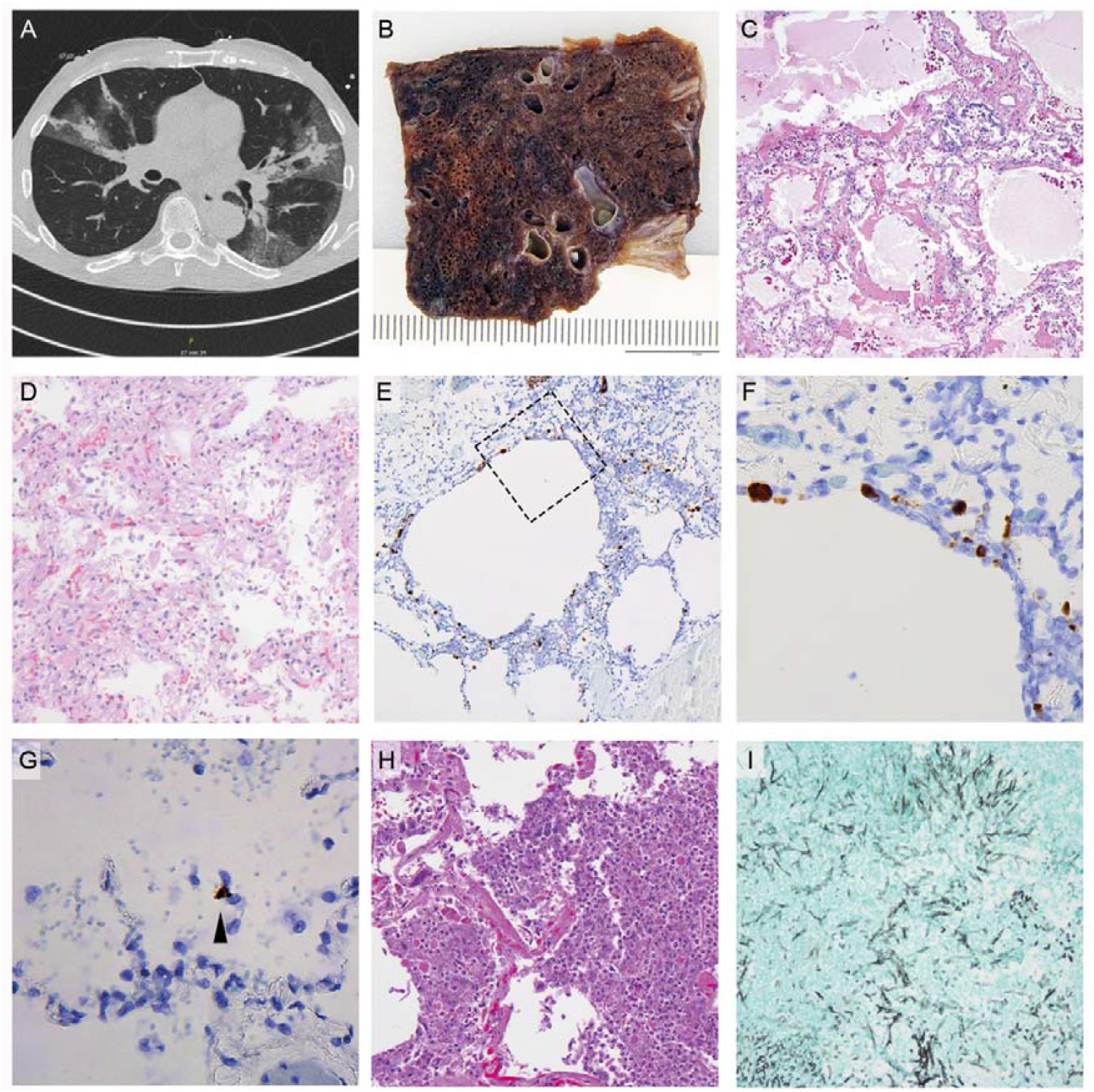
**A**) CT-scan of a COVID-19 pneumonia after single vaccination. **B)** Macroscopic image; formalin-fixed; lung parenchyma is widely destroyed with dark areas of hemorrhage and loss of spongious morphology. **C)** H&E 40x magnification; acute DAD with prominent hyaline membranes. **D)** H&E 200x magnification; organizing DAD with fibroblastic proliferation and loss of alveolar spaces. **E**) RNA-ISH 100x magnification; high viral affection of pneumocytes and probably macrophages around emphysematic alveolar structures **F)** higher magnification of the area in E marked by a square. **G)** RNA-ISH 400x magnification; low viral load with only one affected cell (arrowhead). **H)** H&E 400x magnification; acute bacterial pneumonia with dense aggregates of granulocytes within the alveolar spaces. **I)** Grocott 200x magnification; Aspergilloma of the lung.

**Figure 4.**
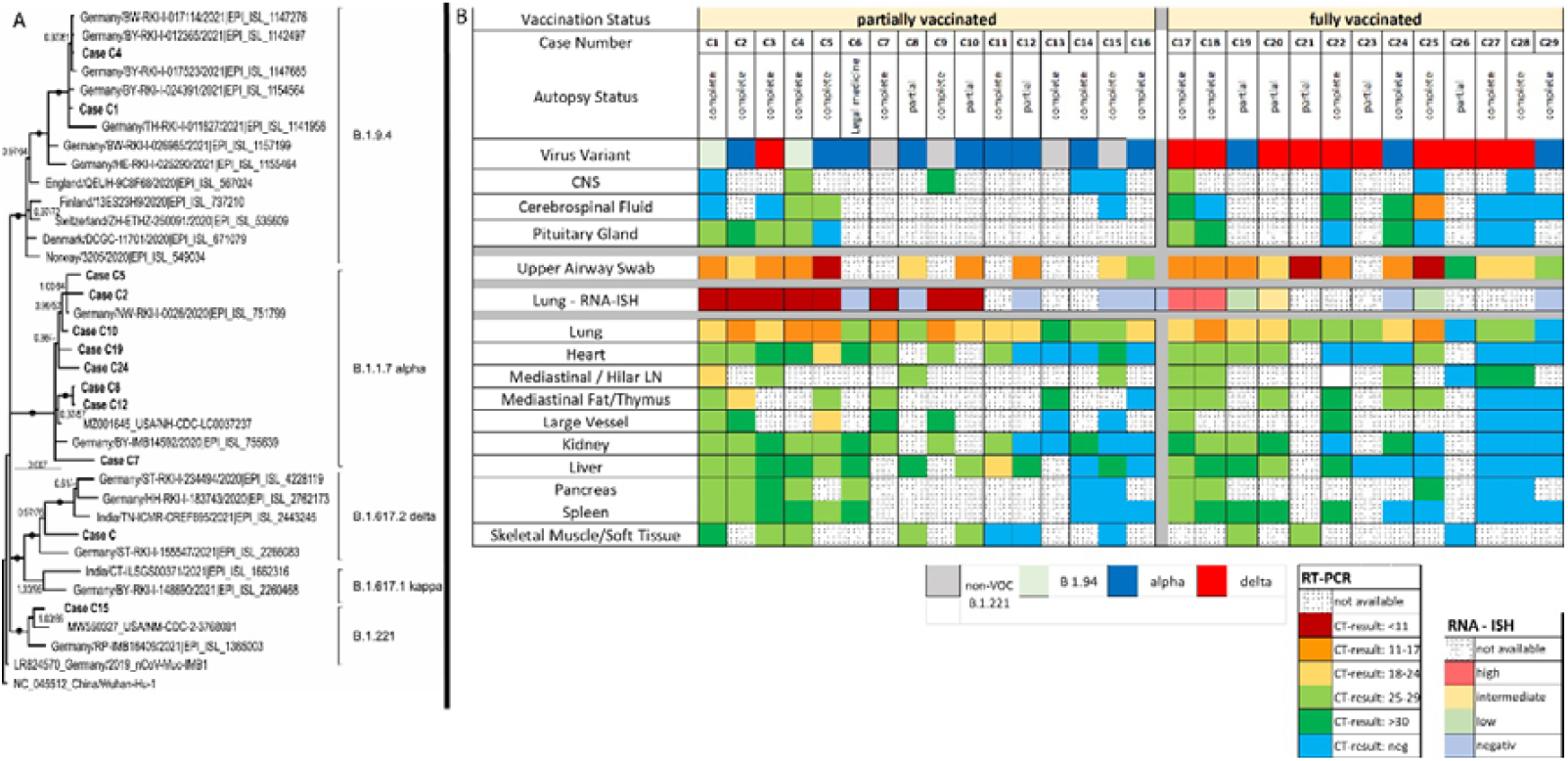
**A)** Phylogenetic tree of SARS-CoV-2 lineages including presented cases. Dots indicate bootstrap values of 1.00/100 (MrBayes/Maximum Likelihood). Support values above 50% are given. **B)** Of note: Due to privacy reasons the cases are not presented in a consecutive manner but are sorted according to the grade of viral dissemination. Autopsy-Status, viral variant lineages and viral affection in different organs by RT-PCR and RNA-ISH (for lungs only). Note: Cases are not sorted in consecutive manner but grade of viral dissemination

In partially vaccinated breakthrough infection patients, the lungs were the most affected organs. with High viral loads could be detected by RT-qPCR, with a median Ct value of 21 (range: 14–31), confirmed by RNA-ISH (Figure 3), which showed a strong correlation with the Ct values (R = 0.819, P < 0.0001) in a semiquantitative analysis (Figure 4). Another remarkable observation in this collection was the high rate of malignancies in their medical history, at 56%. Again, this is considerably higher compared to the completely vaccinated cases (3 vs. 10, 23%; P = 0.130), and the naïve control cohort (30 vs. 112, 21%; P = 0.005).

### Fully Vaccinated Cases

This study comprised 13 fully vaccinated cases. The median time from the last vaccination to a positive test of SARS-CoV-2 was 140 days (range: 28–283). The vaccines applied in the study groups were BNT162b2 (BioNTech) in 11 cases and AZD1222 (AstraZeneca) and CoronaVac (Sinovac) in one case each. The gender ratio was balanced. No correlation between vaccine type and other clinical findings was observed. SARS-CoV-2 VOCs alpha and delta were the only viral variants present (Table 2 and Figures 2 and 4).

According to the WHO classification (33), all but one patient died due to COVID-19. The non-COVID-19 patient suffered from myocardial infarction and renal abscesses. The SARS-CoV-2 RT-qPCR during routine testing on admission was positive, with a very high Ct value of 41. The condition leading directly to death, which does not match the definition of the WHO, was other than COVID-19 (Table 2, Figure 1) at a rate of 31% (4 of 13). The corresponding rates were 25% (4 of 16; P = 1.000) and 8% (11 of 141; P = 0.029) in partially vaccinated cases and nonvaccinated cases, respectively.

The type of infection (“real breakthrough” versus “vaccination failure,” according to Schieffelin et al. (7)) could be classified in 11 of the 13 cases in which serology for SARS-CoV-2 spike-specific antibodies was positive. Five of these cases revealed high anti-spike titers (> 2500 U/ml) while others showed only moderate levels (154 to 407 U/ml). In the remaining two cases, no antibodies against the spike protein were detected (Table 2). According to these results, eight cases were classified as breakthrough infections, whereas two cases represented vaccine failures. In one case, only an asymptomatic infection was obtained. The SARS-CoV-2 nucleocapsid antibody serology revealed negative results (COI < 1) in six of 11 cases, which correlated significantly with the occurrence of a strongly increased or generalized viral dissemination (P = 0.015).

In contrast to the partially vaccinated cases, the viral spread in fully vaccinated cases was restricted to the upper airways and lungs in eight of the 13 cases, whereas viral dissemination throughout the body was seen in five cases. Again, histological changes in the organs were similar to nonvaccinated cases with relevant impairment of the lungs, but only mild changes in other organs, if any. The median RT-PCR Ct value of the lungs was 23 (range: 17–27), similar to the partially vaccinated cases (median 21, range 14–31).

## DISCUSSION

To the best of our knowledge, this is the first series of autopsies of fatal cases of COVID-19 in SARS-CoV-2-vaccinated individuals. The lack of reliable studies and data make it difficult to assess the situation of vaccinated individuals. Therefore, we started to assess viral dissemination in the context of demographic and clinical data to identify potential factors that foster a fatal course of COVID-19 in vaccinees. The aim of this study was to investigate a cohort of 29 fatal COVID-19 cases in vaccinees by collecting all available metadata and by using necropsy, antigen staining and in situ hybridization, RT-qPCR analysis, and whole-genome sequencing for analyzing the course of infection, allowing a substantiated disease and strain characterization.

The focus was on the comparison between partially vaccinated (vaccination interval not completed) and fully vaccinated cases (vaccination interval completed). Moreover, a collection of 141 consecutive cases from nonvaccinated individuals from the Augsburg autopsy series served as controls. Overall, the cases in vaccinees represent about one-third of all deceased in the Augsburg medical center, showing a similar but not identical demographic feature, with a slightly lower proportion of women and a slightly higher age compared to the total collective. All fully vaccinated cases came from the University Medical Center Augsburg, while six of the 16 partially vaccinated cases were contributed by other academic centers.

Given the 303 cases of deceased with COVID-19 during the time of vaccine availability in the Augsburg Center, the 42 (14%) deceased with COVID-19 after vaccination, of which 23 (autopsy rate 55%) are included in this study, indicates that the study cohort is representative.

The University Medical Center Augsburg is the only tertiary medical center in a region with about two million inhabitants. For the city of Augsburg, the rate of completely vaccinated persons is 65% (194,000 persons; status: 4 November 2021) (40). The 16 deceased cases after full vaccination represent a rate of 0.008%, which is considerably low and in line with a large population-based study carried out in Scotland with a rate of 0.007% (41).

The alpha and the delta variants are overrepresented in the groups of the partially and the fully vaccinated cases, respectively. However, this is most likely caused by the course of the pandemic with the alpha variant being most prevalent during a time most people received the first dose of the vaccine just recently and the delta variant that dominates in a phase where the vaccination titers are declining in large parts of the population. A further increase of breakthrough infections can be assumed since the protective effect after vaccination is continuously waning without booster shots and novel strains with immune escape properties are expected.

This study includes two fundamentally different post-vaccination situations, i.e., with partial and full vaccinations. In fully vaccinated cases, the type of infection was classified according to Schieffelin et al. (7), taking so-called “vaccine nonresponders” into account. However, in our study group of fully vaccinated cases, real “breakthrough infections” occurred in the majority of individuals, and only two of nine cases were defined as likely “vaccination failure,” which therefore might play a limited role in lethal infections. For vaccination failure, it has to be further discussed whether it was a primary failure (e.g., nonresponders, application errors, etc.) or loss of vaccination response over time, as recently described in Israel (42). In our study group, based on serological data, a primary failure due to nonresponding, e.g., during steroid treatment, is the most likely cause in both described cases (C17) (C29)(43).

The macroscopic and histomorphological findings in the partially vaccinated deceased were similar to the findings in the nonvaccinated cases. Most patients died due to COVID-19 pneumonia with typical DAD. Superinfections (Tables 1 and 2) occurred at a relatively high frequency (11 of 29), including aspergillosis (four cases). This is considerably more often compared to our previous results (17), but rarer than reported in deceased patients after long-term treatment (44). Other organs generally showed no histological alterations that could be associated with SARS-CoV-2 infection. However, a high rate of viral dissemination within the body was an unanticipated result in this study, which was especially accentuated in the partially vaccinated compared to fully vaccinated cases (11 of 16 vs. five of 13, respectively; P = 0.144). In comparison, such disseminations were previously found in only three of 19 deceased (17). In several cases, RT-qPCR identified the RNA of SARS-CoV-2 in all investigated sample matrices, including cerebrospinal fluid, CNS, and soft tissues. This is in strong contrast to a previously published collection of the Augsburg series of nonvaccinated lethal SARS-CoV-2 infections, in which the frequency of viral dissemination was rare, with a rate of only 16% (three of 19) (17) instead of 69%. In this context, it seems especially important to compare the results of different cohorts within the same analytic system. Other authors have reported results we classify in this study as “disseminated” at high frequencies (45, 46), but use other settings and methods.

Low Ct values of nasopharyngeal swabs and lung samples, the latter with abundant viral detection by RNA-ISH, underline strikingly high viral loads in vaccinated deceased individuals, again with accentuation in partially vaccinated individuals. However, at this point, it must be mentioned that the previous series (17) did not include VOCs. Therefore, it cannot be ruled out that the reported increased viral loads are in part also a consequence of the respective circulating viral variants. However, because we also found this effect in non-VOC vaccinees, and also observed anecdotical restricted dissemination of VOCs including the delta and the gamma variants in non-vaccinees (data not shown), it is probable that the dissemination phenotype observed here is not related to the given variant. A recently published study showed that a single shot of AZD1222 or BNT162b2 showed a relevant effect of protection against infection with SARS-CoV-2 (47). However, this does not equate to complete protection, and individual fatal courses, e.g., also related to preexisting disease conditions, are supported by our data.

Two major contrary theses that could explain this viral spread are 1) the vaccination itself and 2) the constitution of the individual. The first is mediated by antibody-dependent enhancement (ADE) (48-52)), which is known from other viral infections, such as dengue (53), Ebola (54), and HIV (55). In ADE, antibodies do not eliminate the virus or do so only to a reduced extent, but instead promote viral uptake into the host’s cells. Virus-bound IgG is carried into immune cells by Fc-receptor-mediated internalization. The extent to which ADE plays a role in coronavirus infections is unclear. Reports advocating the existence of ADE in coronavirus infections are based on experiments using cell cultures (56, 57) or animal models (58). However, there is currently no evidence for ADE as a relevant mechanism counteracting the protective role of anti-spike protein antibodies generating vaccines in humans. A large study enrolling 20,000 patients receiving COVID-19 convalescent plasma identified no safety concerns (59), which can also be considered a powerful argument against the relevant role of ADE in humans. Currently, no assays or biomarkers have been established to prove ADE in vivo. Immune cell infiltration, including eosinophils indicating an adverse immune reaction, is restricted to T-helper cell-mediated responses and is not related to ADE (60).

Focusing on potential patient-related factors, the immune system is of major interest in the context of failing viral elimination. Both collections in this study are characterized by a high median age and a high rate of potentially immune compromising conditions, such as cancer history (12 individuals), intake of immunosuppressive drugs (three individuals), asplenia (one individual), or decreased immunoglobulin levels (three individuals). One or more of these conditions were found in 69% and 40% of partially and fully vaccinated patients, respectively. A very recent clinical study underlines the role of immune compromission (61).The finding that negative nucleocapsid antibody testing was associated with strongly increased or generalized viral dissemination in fully vaccinated cases (Table 2) further supports the hypothesis that the immune system of these patients was no longer able to elicit a primary response versus the SARS-CoV-2 nucleocapsid protein, while spike-specific antibodies were often present or even boosted to high titers (Table 2). In terms of cancer, a recently published study showed that malignancies are important risk factors for COVID-19, hospitalization, and death (62). One explanation for this finding was the lower rate of seroconversion after vaccination of cancer patients in general as a result of immunosuppression (disease and therapy) (63, 64). The same is true for immunosuppressive antirheumatic drugs (43).

A general limitation of autopsy studies like ours is the rather small case number. In an ongoing pandemic, inhomogeneities regarding the included variants might further weaken the study. Nevertheless, the consecutively collected cases with an appropriate rate can be assumed to be representative enough to draw relevant conclusions.

Overall, this is the first series of fatal courses of COVID-19 after vaccination that was analyzed in detail using a broad range of diagnostic techniques. As a major outcome, it can be concluded that most of the deceased were elderly patients with a high number of comorbidities. Lethal SARS-CoV-2 infection in vaccinated individuals therefore seems to be a very rare event and is mainly connected with a high age and additional underlying factors, such as chronic diseases. A high viral affection, both in terms of the spread within the organism and viral load, together with high rates of immunocompromising conditions, are the most striking findings of this study that were accentuated in cases with an incomplete vaccination status.

## Data Availability

All data produced in the present work are contained in the manuscript

## Acknowledgments

The authors are thankful to Alexandra Martin, Melanie Spörel, Nadine Eismann, Joanna Schmucker, Christian Beul, Lukas Borcherding, Korbinian Krieger, and Stefanie Weber (MD), from Augsburg, G. Crudele, and N. Schalle from Düsseldorf, and Patrick Zitzow for thorough archive work and excellent technical assistance. They further thank Dr. Marlene Lessel (Kaufbeuren), and Eva Exner (MD) (Gesundheitsamt Ostallgäu) for contributing cases.

The authors are particularly thankful to all relatives who gave their consent for the postmortem examination and thus made an invaluable contribution to the research on this new disease.

This work was supported by the German Registry of COVID-19 Autopsies (www.DeRegCOVID.ukaachen.de), funded by the Federal Ministry of Health (ZMVI1-2520COR201), by the Federal Ministry of Education and Research within the framework of the network of university medicine (DEFEAT PANDEMICs, 01KX2021) and the German Federal Ministry of Food and Agriculture through the Federal Office for Agriculture and Food, project ZooSeq, grant number 2819114019.

## References

1. Pascual-Iglesias A, Canton J, Ortega-Prieto AM, Jimenez-Guardeno JM, Regla-Nava JA. An Overview of Vaccines against SARS-CoV-2 in the COVID-19 Pandemic Era. Pathogens. 2021;10(8).

2. Jackson LA, Roberts PC, Graham BS. A SARS-CoV-2 mRNA Vaccine - Preliminary Report. Reply. N Engl J Med. 2020;383(12):1191–2.

3. Polack FP, Thomas SJ, Kitchin N, Absalon J, Gurtman A, Lockhart S, et al. Safety and Efficacy of the BNT162b2 mRNA Covid-19 Vaccine. N Engl J Med. 2020;383(27):2603–15.

4. Sadoff J, Gray G, Vandebosch A, Cardenas V, Shukarev G, Grinsztejn B, et al. Safety and Efficacy of Single-Dose Ad26.COV2.S Vaccine against Covid-19. N Engl J Med. 2021;384(23):2187–201.

5. Voysey M, Clemens SAC, Madhi SA, Weckx LY, Folegatti PM, Aley PK, et al. Safety and efficacy of the ChAdOx1 nCoV-19 vaccine (AZD1222) against SARS-CoV-2: an interim analysis of four randomised controlled trials in Brazil, South Africa, and the UK. Lancet. 2021;397(10269):99–111.

6. Hacisuleyman E, Hale C, Saito Y, Blachere NE, Bergh M, Conlon EG, et al. Vaccine Breakthrough Infections with SARS-CoV-2 Variants. N Engl J Med. 2021;384(23):2212–8.

7. Schieffelin JS, Norton EB, Kolls JK. What should define a SARS-CoV-2 “breakthrough” infection? J Clin Invest. 2021;131(12).

8. Lustig Y, Sapir E, Regev-Yochay G, Cohen C, Fluss R, Olmer L, et al. BNT162b2 COVID-19 vaccine and correlates of humoral immune responses and dynamics: a prospective, single-centre, longitudinal cohort study in health-care workers. Lancet Respir Med. 2021;9(9):999–1009.

9. Robert-Koch-Institut 2021;Pageshttps://www.rki.de/DE/Content/InfAZ/N/Neuartiges_Coronavirus/Situationsberichte/Wochenbericht/Wochenberichte_Tab.html;jsessionid=7A9EF1DE85D2947A7C499E14A1C5E07C.internet092?nn=13490888 on 30.10.2021 2021.

10. Tenforde MW, Olson SM, Self WH, Talbot HK, Lindsell CJ, Steingrub JS, et al. Effectiveness of Pfizer-BioNTech and Moderna Vaccines Against COVID-19 Among Hospitalized Adults Aged >/=65 Years -United States, January-March 2021. MMWR Morb Mortal Wkly Rep. 2021;70(18):674–9.

11. Lutrick K, Ellingson KD, Baccam Z, Rivers P, Beitel S, Parker J, et al. COVID-19 Infection, Reinfection, and Vaccine Effectiveness in a Prospective Cohort of Arizona Frontline/Essential Workers: The AZ HEROES Research Protocol. JMIR Res Protoc. 2021.

12. Thompson MG, Burgess JL, Naleway AL, Tyner HL, Yoon SK, Meece J, et al. Interim Estimates of Vaccine Effectiveness of BNT162b2 and mRNA-1273 COVID-19 Vaccines in Preventing SARS-CoV-2 Infection Among Health Care Personnel, First Responders, and Other Essential and Frontline Workers - Eight U.S. Locations, December 2020-March 2021. MMWR Morb Mortal Wkly Rep. 2021;70(13):495–500.

13. Planas D, Bruel T, Grzelak L, Guivel-Benhassine F, Staropoli I, Porrot F, et al. Sensitivity of infectious SARS-CoV-2 B.1.1.7 and B.1.351 variants to neutralizing antibodies. Nat Med. 2021;27(5):917–24.

14. Planas D, Veyer D, Baidaliuk A, Staropoli I, Guivel-Benhassine F, Rajah MM, et al. Reduced sensitivity of SARS-CoV-2 variant Delta to antibody neutralization. Nature. 2021;596(7871):276–80.

15. Tregoning JS, Flight KE, Higham SL, Wang Z, Pierce BF. Progress of the COVID-19 vaccine effort: viruses, vaccines and variants versus efficacy, effectiveness and escape. Nat Rev Immunol. 2021;21(10):626–36.

16. Harvey WT, Carabelli AM, Jackson B, Gupta RK, Thomson EC, Harrison EM, et al. SARS-CoV-2 variants, spike mutations and immune escape. Nat Rev Microbiol. 2021;19(7):409–24.

17. Hirschbühl K, Dintner S, Beer M, Wylezich C, Schlegel J, Delbridge C, et al. Viral mapping in COVID-19 deceased in the Augsburg autopsy series of the first wave: A multiorgan and multimethodological approach. PLoS One. 2021;16(7):e0254872.

18. Massoth LR, Desai N, Szabolcs A, Harris CK, Neyaz A, Crotty R, et al. Comparison of RNA In Situ Hybridization and Immunohistochemistry Techniques for the Detection and Localization of SARS-CoV-2 in Human Tissues. Am J Surg Pathol. 2021;45(1):14–24.

19. Calabrese F, Pezzuto F, Fortarezza F, Hofman P, Kern I, Panizo A, et al. Pulmonary pathology and COVID-19: lessons from autopsy. The experience of European Pulmonary Pathologists. Virchows Arch. 2020;477(3):359–72.

20. Pomara C, Li Volti G, Cappello F. COVID-19 Deaths: Are We Sure It Is Pneumonia? Please, Autopsy, Autopsy, Autopsy! J Clin Med. 2020;9(5).

21. Salerno M, Sessa F, Piscopo A, Montana A, Torrisi M, Patanè F, et al. No Autopsies on COVID-19 Deaths: A Missed Opportunity and the Lockdown of Science. J Clin Med. 2020;9(5).

22. Satturwar S, Fowkes M, Farver C, Wilson AM, Eccher A, Girolami I, et al. Postmortem Findings Associated With SARS-CoV-2: Systematic Review and Meta-analysis. Am J Surg Pathol. 2021.

23. Bösmüller H, Traxler S, Bitzer M, Haberle H, Raiser W, Nann D, et al. The evolution of pulmonary pathology in fatal COVID-19 disease: an autopsy study with clinical correlation. Virchows Arch. 2020;477(3):349–57.

24. Wichmann D, Sperhake JP, Lutgehetmann M, Steurer S, Edler C, Heinemann A, et al. Autopsy Findings and Venous Thromboembolism in Patients With COVID-19: A Prospective Cohort Study. Ann Intern Med. 2020;173(4):268–77.

25. Menter T, Haslbauer JD, Nienhold R, Savic S, Hopfer H, Deigendesch N, et al. Postmortem examination of COVID-19 patients reveals diffuse alveolar damage with severe capillary congestion and variegated findings in lungs and other organs suggesting vascular dysfunction. Histopathology. 2020.

26. Hanley B, Naresh KN, Roufosse C, Nicholson AG, Weir J, Cooke GS, et al. Histopathological findings and viral tropism in UK patients with severe fatal COVID-19: a post-mortem study. Lancet Microbe. 2020.

27. Ackermann M, Verleden SE, Kuehnel M, Haverich A, Welte T, Laenger F, et al. Pulmonary Vascular Endothelialitis, Thrombosis, and Angiogenesis in Covid-19. N Engl J Med. 2020;383(2):120–8.

28. Varga Z, Flammer AJ, Steiger P, Haberecker M, Andermatt R, Zinkernagel AS, et al. Endothelial cell infection and endotheliitis in COVID-19. Lancet. 2020;395(10234):1417–8.

29. Püschel K, Sperhake JP. Corona deaths in Hamburg, Germany. Int J Legal Med. 2020;134(4):1267–9.

30. Lax SF, Skok K, Zechner P, Kessler HH, Kaufmann N, Koelblinger C, et al. Pulmonary Arterial Thrombosis in COVID-19 With Fatal Outcome : Results From a Prospective, Single-Center, Clinicopathologic Case Series. Ann Intern Med. 2020;173(5):350–61.

31. Hansen T, Titze U, Kulamadayil-Heidenreich NSA, Glombitza S, Tebbe JJ, Rocken C, et al. First case of postmortem study in a patient vaccinated against SARS-CoV-2. Int J Infect Dis. 2021;107:172–5.

32. Berezowska S, Boor P, Jonigk D, Tischler V. Update on thoracic pathology 2021-report of the working group thoracic pathology of the German Society of Pathology. Der Pathologe. 2021.

33. WHO 2020;Pageshttps://www.who.int/classifications/icd/Guidelines_Cause_of_Death_COVID-19.pdf on 05.04.2021 2021.

34. Wylezich C, Papa A, Beer M, Hoper D. A Versatile Sample Processing Workflow for Metagenomic Pathogen Detection. Sci Rep. 2018;8(1):13108.

35. Wylezich C, Calvelage S, Schlottau K, Ziegler U, Pohlmann A, Hoper D, et al. Next-generation diagnostics: virus capture facilitates a sensitive viral diagnosis for epizootic and zoonotic pathogens including SARS-CoV-2. Microbiome. 2021;9(1):51.

36. Rambaut A, Holmes EC, O’Toole A, Hill V, McCrone JT, Ruis C, et al. A dynamic nomenclature proposal for SARS-CoV-2 lineages to assist genomic epidemiology. Nat Microbiol. 2020;5(11):1403–7.

37. Katoh K, Standley DM. MAFFT multiple sequence alignment software version 7: improvements in performance and usability. Mol Biol Evol. 2013;30(4):772–80.

38. Guindon S, Dufayard JF, Lefort V, Anisimova M, Hordijk W, Gascuel O. New algorithms and methods to estimate maximum-likelihood phylogenies: assessing the performance of PhyML 3.0. Syst Biol. 2010;59(3):307–21.

39. Ronquist F, Huelsenbeck JP. MrBayes 3: Bayesian phylogenetic inference under mixed models. Bioinformatics. 2003;19(12):1572–4.

40. Augsburg Co 2021;Pageshttps://www.augsburg.de/umwelt-soziales/gesundheit/coronavirus/fallzahlen on 18.11.2021 2021.

41. Grange Z, Buelo A, Sullivan C, Moore E, Agrawal U, Boukhari K, et al. Characteristics and risk of COVID-19-related death in fully vaccinated people in Scotland. The Lancet. 2021.

42. Goldberg Y, Mandel M, Bar-On YM, Bodenheimer O, Freedman L, Haas EJ, et al. Waning Immunity after the BNT162b2 Vaccine in Israel. N Engl J Med. 2021.

43. Friedman MA, Curtis JR, Winthrop KL. Impact of disease-modifying antirheumatic drugs on vaccine immunogenicity in patients with inflammatory rheumatic and musculoskeletal diseases. Ann Rheum Dis. 2021;80(10):1255–65.

44. Evert K, Dienemann T, Brochhausen C, Lunz D, Lubnow M, Ritzka M, et al. Autopsy findings after long-term treatment of COVID-19 patients with microbiological correlation. Virchows Arch. 2021:1–12.

45. Remmelink M, De Mendonça R, D’Haene N, De Clercq S, Verocq C, Lebrun L, et al. Unspecific post-mortem findings despite multiorgan viral spread in COVID-19 patients. Crit Care. 2020;24(1):495.

46. Wong DWL, Klinkhammer BM, Djudjaj S, Villwock S, Timm MC, Buhl EM, et al. Multisystemic Cellular Tropism of SARS-CoV-2 in Autopsies of COVID-19 Patients. Cells. 2021;10(8).

47. Shrotri M, Krutikov M, Palmer T, Giddings R, Azmi B, Subbarao S, et al. Vaccine effectiveness of the first dose of ChAdOx1 nCoV-19 and BNT162b2 against SARS-CoV-2 infection in residents of long-term care facilities in England (VIVALDI): a prospective cohort study. Lancet Infect Dis. 2021;21(11):1529–38.

48. Sanchez-Zuno GA, Matuz-Flores MG, Gonzalez-Estevez G, Nicoletti F, Turrubiates-Hernandez FJ, Mangano K, et al. A review: Antibody-dependent enhancement in COVID-19: The not so friendly side of antibodies. Int J Immunopathol Pharmacol. 2021;35:20587384211050199.

49. Maemura T, Kuroda M, Armbrust T, Yamayoshi S, Halfmann PJ, Kawaoka Y. Antibody-Dependent Enhancement of SARS-CoV-2 Infection Is Mediated by the IgG Receptors FcgammaRIIA and FcgammaRIIIA but Does Not Contribute to Aberrant Cytokine Production by Macrophages. mBio. 2021;12(5):e0198721.

50. Lee WS, Wheatley AK, Kent SJ, DeKosky BJ. Antibody-dependent enhancement and SARS-CoV-2 vaccines and therapies. Nat Microbiol. 2020;5(10):1185–91.

51. Bournazos S, Gupta A, Ravetch JV. The role of IgG Fc receptors in antibody-dependent enhancement. Nat Rev Immunol. 2020;20(10):633–43.

52. Arvin AM, Fink K, Schmid MA, Cathcart A, Spreafico R, Havenar-Daughton C, et al. A perspective on potential antibody-dependent enhancement of SARS-CoV-2. Nature. 2020;584(7821):353–63.

53. Halstead SB, Chow JS, Marchette NJ. Immunological enhancement of dengue virus replication. Nat New Biol. 1973;243(122):24–6.

54. Takada A, Feldmann H, Ksiazek TG, Kawaoka Y. Antibody-dependent enhancement of Ebola virus infection. J Virol. 2003;77(13):7539–44.

55. Laurence J, Saunders A, Early E, Salmon JE. Human immunodeficiency virus infection of monocytes: relationship to Fc-gamma receptors and antibody-dependent viral enhancement. Immunology. 1990;70(3):338–43.

56. Wang SF, Tseng SP, Yen CH, Yang JY, Tsao CH, Shen CW, et al. Antibody-dependent SARS coronavirus infection is mediated by antibodies against spike proteins. Biochem Biophys Res Commun. 2014;451(2):208–14.

57. Jaume M, Yip MS, Cheung CY, Leung HL, Li PH, Kien F, et al. Anti-severe acute respiratory syndrome coronavirus spike antibodies trigger infection of human immune cells via a pH- and cysteine protease-independent FcgammaR pathway. J Virol. 2011;85(20):10582–97.

58. Hoffmann D, Corleis B, Rauch S, Roth N, Muhe J, Halwe NJ, et al. CVnCoV and CV2CoV protect human ACE2 transgenic mice from ancestral B BavPat1 and emerging B.1.351 SARS-CoV-2. Nat Commun. 2021;12(1):4048.

59. Joyner MJ, Bruno KA, Klassen SA, Kunze KL, Johnson PW, Lesser ER, et al. Safety Update: COVID-19 Convalescent Plasma in 20,000 Hospitalized Patients. Mayo Clin Proc. 2020;95(9):1888–97.

60. Hotez PJ, Corry DB, Bottazzi ME. COVID-19 vaccine design: the Janus face of immune enhancement. Nat Rev Immunol. 2020;20(6):347–8.

61. Di Fusco M, Moran MM, Cane A, Curcio D, Khan F, Malhotra D, et al. Evaluation of COVID-19 vaccine breakthrough infections among immunocompromised patients fully vaccinated with BNT162b2. J Med Econ. 2021;24(1):1248–60.

62. Roel E, Pistillo A, Recalde M, Fernández-Bertolín S, Aragón M, Soerjomataram I, et al. Cancer and the risk of coronavirus disease 2019 diagnosis, hospitalisation and death: A population-based multistate cohort study including 4[]618[]377 adults in Catalonia, Spain. Int J Cancer. 2021.

63. Thakkar A, Gonzalez-Lugo JD, Goradia N, Gali R, Shapiro LC, Pradhan K, et al. Seroconversion rates following COVID-19 vaccination among patients with cancer. Cancer Cell. 2021;39(8):1081–90 e2.

64. Monin-Aldama L, Laing AG, Muñoz-Ruiz M, McKenzie DR, del Molino del Barrio I, Alaguthurai T, et al. Interim results of the safety and immune-efficacy of 1 versus 2 doses of COVID-19 vaccine BNT162b2 for cancer patients in the context of the UK vaccine priority guidelines. medRxiv. 2021.

